# Treatment response biomarkers in early Alzheimer’s disease: longitudinal trajectories, sample size estimates, and the impact of progression variability

**DOI:** 10.64898/2026.08.27.26361425

**Authors:** Marlies Oosthoek, Alida Leistra, Yanaika S. Hok-A-Hin, Michael W. T. Tanck, Tomohiro Okuda, Lisanne in ‘t Veld, Azzam Aladdin, Pieter van Bokhoven, Betty M. Tijms, Roos J. Jutten, Philip Scheltens, Everard G.B. Vijverberg, Charlotte E. Teunissen, Lisa Vermunt

## Abstract

**Background:** Fluid biomarkers enable the demonstration of the biological effects of novel therapies in Alzheimer’s disease (AD). However, longitudinal biomarker data are sparse and sample size calculations for fluid biomarkers are often lacking. Here, we provided longitudinal CSF and plasma AD biomarkers measured in samples collected in a placebo arm in a 1.5-year phase 2b trial, allowing us to study natural trajectories, required sample sizes and heterogeneity in early AD clinical trials.

**Methods:** We studied individuals from the placebo group (MCI due to AD (n=65) and AD dementia (n=41)) of the T-817MA trial (NCT04191486) with positive CSF AD biomarkers (mean age=69±7 years, Female=63%). Longitudinal biomarker changes in CSF (Aβ42, Aβ40, Aβ42/40, pTau181, pTau217, NFL, tTau, YKL40, NRGN, ABL1, CHIT1, CLEC5A, ITGB2, MMP10, SDC4, SPON2, THBD) and plasma biomarkers (Aβ42, Aβ40, Aβ42/40, pTau181, pTau217, NFL, GFAP) were analyzed with linear mixed-effect models. Required sample size estimates for predefined treatment effects were generated. Lastly, we investigated the influence of between person variability in biomarker change by simulating a randomized clinical trial (1:1) 10000 times, and assessed the group differences at 1.5 years.

**Findings:** Fourteen biomarkers changed over time, with the largest annual changes observed for plasma pTau217 (+9.8%), CSF MMP10 (+7.1%), and CSF NFL (+6.9%), and CSF Aβ40 by (−4.0%), CSF pTau217 (−3.0%), and CSF NRGN (−2.5%). To show a 30% change, similar to biomarker effects of approved AD drugs, almost all markers required less than 45 patients per trial arm. To reach normalized levels, established CSF markers required lower sample sizes than plasma markers. The effects of heterogeneity over time were approximately twice as large in plasma compared to CSF.

**Interpretation:** These findings offer insights into the biomarker trajectories and power in early AD, supporting more informed endpoint selection and forming a frame of reference for the interpretation of treatment effects in clinical trials.

## Introduction

Alzheimer’s disease (AD) is a multifaceted disease and new disease-modifying therapies (DMTs) that investigate a wide range of targets are under development^1–4^. Fluid biomarkers offer the potential to establish target engagement or disease modification in a short time frame and measure multiple biological processes within one sample^3^. However, there is a lack of data to inform the selection of fluid biomarker endpoints. Therefore, establishing the biomarker trajectories and estimating the required sample sizes of established and emerging CSF and plasma biomarkers can provide crucial input for the design and interpretation of future studies.

Core AD pathological hallmarks are reflected by diagnostic markers in CSF and plasma of amyloid and tau isoforms^5–14^. Neurofilament light (NFL) can be measured in CSF and plasma and is an overall indicator of neurodegeneration in AD^15–17^. Plasma glial fibrillary acidic protein (GFAP) and CSF chitinase-3-like protein 1 (YKL40) are glial markers of neuroinflammation^18,19^ and CSF neurogranin (NRGN) is a marker for synaptic dysfunction^20^. Observational studies have shown that over a short period of 6 months the core CSF biomarkers (Aβ42, pTau181 and tTau) and YKL40 were stable^21,22^. With longer follow up, CSF Aβ42 levels stabilized or decreased^23–25^. In the MCI stage, CSF pTau181 and CSF tTau levels increased over time, while in the dementia stage, the overall pattern showed that CSF tau levels stabilized or decreased, with the most clearly defined decreases in CSF pTau181 levels^23–30^. CSF NRGN levels also increased over time in the MCI stage and stabilized or decreased in the AD dementia stage^24,31^. CSF YKL40 levels increased in the MCI stage and tended to continue to increase in the dementia stage^24,26,32^. CSF NFL levels increased by 4-5% per year, but no relationship with clinical stage could be detected^26,31^. Longitudinal studies in plasma biomarkers showed that pTau181, pTau217, GFAP and NFL levels generally increased or were stable over time in the MCI and AD dementia stages^14,33–37^. These trajectories could differ in a clinical trial setting, with a selected population and a structured design for repeated sampling and total follow-up^38^. In addition, a recently developed AD dementia panel has been proposed as treatment response markers for future drugs that target novel mechanisms in AD^1,3,4,39,40^. These markers include ABL1 (protein phosphorylation), SDC4 (exosome assembly), MMP10 (cytoskeletal remodeling), THBD (vascular function), ITGB2, CLEC5A, CHIT1, and SPON2 (immune system)^1,41–43^. However, longitudinal measurements are needed to support their utility as treatment response biomarkers.

Therefore, the aim of this study was to investigate natural disease progression in a trial setting and determine how disease heterogeneity can affect this progression and subsequently required sample sizes and clinical trial interpretation. We measured established and novel plasma and CSF biomarkers to 1) examine biomarker trajectories, 2) calculate sample size estimates, and 3) assess the effects of heterogeneity of biomarker changes over time in the placebo group of a well-conducted phase 2 clinical trial. The trial included a prodromal to mild AD population. CSF and plasma was drawn at baseline, 1 year and 1.5 years. This comprehensive approach allowed us to gain insights into the trajectory and within-person stability of the novel biomarkers, establish and compare the power for each of the biomarkers, and provide a frame of reference for the interpretation of biomarker results of AD clinical trials.

## Methods

### Patient selection

Participants were enrolled in the T-817MA clinical trial (NCT04191486) that was conducted between December 2019 and March 2023^44,45^. Participants from the placebo group (n=108) with CSF and/or plasma biomarkers available for established an novel biomarkers were included in the study (n=106). Sample availability at 1 year and 1.5 year follow-up was 84 (79%) and 83 (78%) for CSF and 102 (96%) and 100 (94%) for plasma, respectively. Participants were enrolled throughout the EU; in Germany, the Netherlands, the UK, Czechia, Spain, Ireland, and Hungary. The main inclusion criteria of the clinical trial were: a diagnosis of MCI due to AD or mild AD dementia according to NIA-AA diagnostic criteria^46^ with an MMSE of 24 to 30 (inclusive) and the CSF profile was consistent with AD (Elecsys Aβ42 ≤1000 pg/mL, pTau181 ≥19 pg/mL). Main exclusion criteria were: contraindications to a lumbar punction or MRI, a recent MRI scan (<2 years) showing pathology inconsistent with AD, and memantine usage or recent initiation (<3 months) of an acetylcholinesterase inhibitor. The trial was conducted in accordance with ICH guidelines and the ethical principles of the Declaration of Helsinki. The trial was approved by an independent local ethical committee at each participating country/center. All study participants or representatives provided written informed consent prior to performing any study related procedures.

### CSF and plasma draw and biomarker analysis

CSF was obtained by lumbar puncture and collected in polypropylene tubes. Samples were centrifuged 2000 x g for 10 minutes at room temperature and stored at −80 °C until analysis. EDTA plasma was obtained through venipuncture. Samples were centrifuged at 1800 x g for 10 minutes at room temperature. Samples were stored at −80 °C until analysis.

Prior to analyses, samples were thawed and centrifuged at 10000 x g for 10 minutes. The following biomarkers were measured in plasma on single molecule array (SIMOA) HD-X analyzer: Aβ40, Aβ42, NFL, GFAP (mono, the Neurology 4-plex E Advantage kit, Quanterix, USA), pTau217 (duplo, ALZpath CARe Advantage kit, Quanterix, USA), and pTau181 (duplo, Advantage V2 kit, Quanterix, USA). The following biomarkers were measured in CSF: Aβ42, tTau, pTau181 (Cobas Pro 2, Elecsys, Roche Diagnostics GmbH, Germany), NFL (mono, SIMOA HD-X V2 Advantage kit, Quanterix, USA), pTau217 (duplo, SIMOA HD_X ALZpath CARe Advantage kit, Quanterix, USA), YKL40 (duplo, ELISA MicroVue Bone YKL40, Quidel, USA), NRGN (mono, ELISA, Euroimmune, Germany), and Aβ40 (Lumipulse G600 Fujirebio, Belgium). Calibration curves and kit QC samples were included in all measurements and were used to assure stable assay performance (<15% CV). All biomarkers were measured in pg/mL, except YKL40, which was measured in ng/mL.

### Emerging CSF markers multi-dementia panel

Recently, we developed a quantitative custom proximity extension dementia-panel for emerging dementia CSF biomarkers, of which eight that were related to an AD diagnosis were included in this study: ABL1, SDC4, MMP10, ITGB2, CLEC5A, THBD, SPON2, and CHIT1^1^. This panel relies on the proximity extension assay (PEA) technology (Olink Proteomics, Uppsala, Sweden) and measurements were performed on the Signature Q100 Olink platform. Samples were randomized across plates, all timepoints of one participant were put sequentially on the same plate to minimize within person analytical variation.

All plates included four QC samples, one negative control sample, and three single-point calibrators. The QC samples were used to calculate between and within plate coefficients of variation (CVs). All CV% between and within plates were below the 20%, except for ABL1 (38%). However, the QC samples were relatively close to the lower limit of quantification (LLOQ), inflating the CV%. For each protein, the manufacturer established a 24-point standard curve using four-parameter logistic curve fitting. Median values of the single-point calibrators were then applied to align the measurements to this predefined calibration curve. Limits of quantification (LOQ) were established during the development process by the manufacturer and were defined as three standard deviations above the background. LOQ filtering of 85% was applied. Of the eight proteins included in the manuscript, 94% of the samples was above the LOQ. Protein levels were reported in pg/mL. CHIT1 biomarker levels can be affected by rare splice variants, leading to undetectable levels in a small subgroup of individuals. We included these in the original analysis of the 106 participants, four had an SNP resulting in values below LOQ.

### Statistical analysis

Statistical analyses were performed using RStudio version 4.4.3. Demographics were summarized and compared between the MCI due to AD and AD dementia group and between the baseline patients and patients lost to follow-up (LTFU) with t-tests (age, MMSE), Wilcoxon tests (biomarkers), and Chi-square tests (sex and APOE4 carriership).

Longitudinal trajectories for each of the biomarkers were assessed using linear mixed-effects models (Figure 1). The measurements were log transformed to meet homoscedasticity, normality of residuals, and normality of random effects criteria. Time was included as the continuous independent variable, and baseline age and sex were included as covariates. The model included random subject intercepts allowing for between individual variability in biomarker levels at baseline. Random slopes were not included as the models did not converge when they were added. Effects of disease stage (MCI due to AD or mild AD dementia) were investigated by adding an interaction of diagnosis with time to the model. FDR-multiple testing correction was applied to the models and were reported as q-values. To obtain insight in how the within-person variability over time compared to the between-person variability, we calculated the intraclass correlation coefficient (ICC). ICC values were derived from the model as a ratio of the between-participant variation to the total variance (between-participant + residual within participant variance)^47,48^. Biomarkers were classified as having poor (ICC<0.5), moderate (ICC 0.50 – 0.75), good (ICC 0.75 – 0.90), or excellent (ICC > 0.90) longitudinal reliability^49^. ICC values were compared with the Wilcoxon signed rank test. We repeated all analyses without 4 individuals that likely carry a genetic splice variant that lead to low CHIT1 detection to exclude an effect of these low outliers on the overall conclusions^50^. To further characterize the pattern of biomarker changes, the difference between the baseline and end of treatment log values (the log ratio) was calculated per individual. For each pair of biomarkers, the individual log ratios were correlated using Pearson correlation with FDR multiple testing correction (q-value) to identify protein clusters that followed a similar pattern. Correlations were visualized using a correlation heatmap where biomarkers that correlated together formed a cluster.

**Figure 1.**
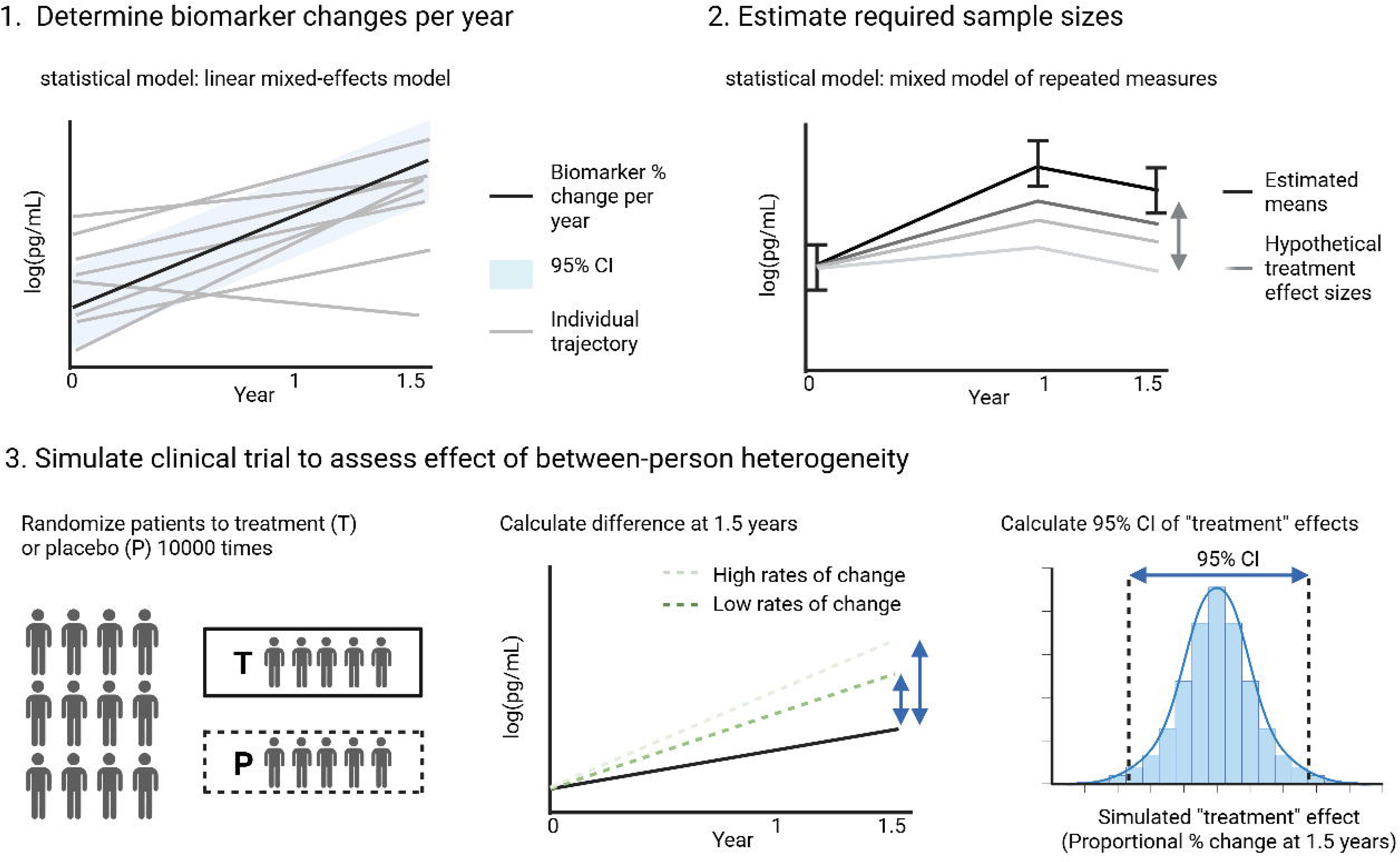
Schematic overview of methods.

Next, we determined minimum required sample sizes per biomarker to detect a range of hypothetical treatment effects (Figure 1). The mixed-model of repeated measures (MMRM), a standard analyses method to determine treatment effects in clinical trials, was used to calculate estimated marginal means per timepoint. The MMRM model does not fit a slope and allows for positive and negative directionality between timepoints, making it suited for clinical trial analysis where treatment effects are not assumed to be linear or follow a predefined direction. Log transformed values of baseline, year 1, and year 1.5 were used as input. Sample sizes were estimated using the longpower package in R under the assumption of 80% power, an expected drop-out rate of 15% at year 1 and 20% after 1.5 years, an unstructured correlation structure, and a significance level of p<0.05. Sample sizes were calculated for a placebo-controlled two-arm trial (1:1) of 1.5 years. To calculate the sample sizes, an expected effect size is required as input. We chose a range of relevant effect sizes based on: 1) relative difference to placebo at end-of-treatment (5%, 10%, 20%, 30%); 2) the degree of normalization towards control levels (10%, 30%, 50%, 100%); 3) change in rate of change for the markers that showed a significant increase in abnormality during the trial period (50% less change, and stabilization). To calculate the effect sizes that aligned with normalization of AD biomarker levels, we used the effect sizes comparing AD and controls available from the biomarker meta-analyses on Alzforum (CSF Aβ40, Aβ42, pTau181, pTau217, tTau, NFL, YKL40, NRGN, plasma Aβ40, Aβ42, pTau181, pTau217, GFAP, and NFL) and from the literature (CSF Aβ42/40, ABL1, CHIT1, CLECL5A, ITGB2, MMP10, SDC4, SPON2, THBD and plasma Aβ42/40,)^1,15,51–54^.

Lastly, we adapted a previously described approach to simulate the possible effects of oversampling fast or slow progressors, to examine to which extend heterogeneity in biomarker changes could affect presumed treatment effects (Figure 1)^55^. For each marker, the previously calculated log ratios were used as input for the simulations. We randomly assigned patients to a treatment or placebo group (1:1) without applying any treatment effect to simulate a clinical trial and calculated the observed difference (log) between the groups: placebo and treatment. We repeated the simulation 10000 times to calculate a 95% confidence interval (CI) of the observed difference between the treatment and placebo group. To facilitate interpretation, the 95% CIs were exponentiated and expressed in percentages. A wider 95% CI indicates that chance alone could mimic a larger apparent treatment effect, which increases the risk of false positives. Simulations were performed allowing resampling and assuming that the trial included 200 participants in total. These simulations were repeated per clinical group to assess the effect disease stage (MCI and mild AD dementia; n=200) and for different sample sizes per group reflecting clinical trial phases (n=50 for phase 1B/2A, n=200 for phase 2B, or n=2000 for phase 3).

## Results

### Participant description

We assessed CSF and plasma samples from 106 participants enrolled in the placebo group of the T-817MA trial of which 61% had MCI due to AD and 39% had mild AD dementia (Table 1). The mean age was 69 years (±7 years) and 63% were women. Comparing the clinical groups, the mild AD dementia group contained more females (73% vs 57%) compared to the MCI group. MMSE scores were lower in the 6 participants lost to follow-up (LTFU) in plasma (Supplemental Table 1). There were no additional differences between the participants LTFU and participants who completed the 1.5 years.

**Table 1.** Baseline cohort characteristics. Mean (SD) unless otherwise specified. Numerical measurements were compared between the MCI and mild AD dementia group using t-tests (age and MMSE) and Wilcoxon signed rank tests (biomarkers). Sex and APOE4 carriership were compared with the chi-square test. Abbreviations: MCI mild cognitive impairment, AD Alzheimer’s disease, MMSE mini-mental state examination, CSF cerebrospinal fluid * p<0.05, ** p<0.01

|  | **Total population**<br>**(N=106)** | **MCI due to AD**<br>**(N=65)** | **Mild AD dementia**<br>**(N=41)** |
| --- | --- | --- | --- |
| **Age, years** | 69 (7) | 69 (7) | 68 (8) |
| **Sex female, N (%)** | 67 (63%) | 37 (57%) | 30 (73%) |
| **MMSE, score** | 27 (1.8) | 27 (1.7)** | 26 (1.8)** |
| **APOE E4 carriers, N(%)** | 70 (66%) | 46 (71%) | 24 (59%) |
| **CSF Aβ42/40 ratio** | 0.05 (0.01) | 0.05 (0.01) | 0.05 (0.01) |
| **CSF pTau181, pg/mL** | 38 (15) | 35 (12) | 42 (19) |
| **CSF tTau, pg/mL** | 374 (138) | 352 (112) | 411 (168) |
| **CSF per visit, N (%)** |  |  |  |
| Baseline | 106 (100%) | 65 (100%) | 41 (100%) |
| Year 1 | 85 (80%) | 51 (79%) | 34 (83%) |
| Year 1.5 | 85 (80%) | 50 (77%) | 35 (85%) |
| **Plasma per visit, N (%)** |  |  |  |
| Baseline | 105 (99%) | 64 (99%) | 41 (100%) |
| Year 1 | 102 (96%) | 62 (95%) | 40 (98%) |
| Year 1.5 | 100 (94%) | 60 (92%) | 40 (98%) |

### Biomarker changes over time during the 1.5 year clinical trial

To investigate the natural disease course for all biomarkers, we first investigated how the biomarkers changed with disease progression over 1.5 years (Table 2; Supplemental Figure 1). Of the 17 CSF biomarkers, 10 significantly changed over time, seven of which increased and three decreased. We found a yearly increase in the following CSF biomarker levels: CSF Aβ42/40 ratio of 2.3% (p=0.0373, q=0.0704), CSF NFL of 6.9% (p<0.0001, q=0.0003), CSF YKL40 of 1.4% (p=0.03821, q=0.0704), CSF SDC4 of 2.2% (p=0.0113, q=0.0311), CSF MMP10 of 7.1% (p<0.0001, q<0.0001), CSF THBD of 4.1% (p=0.0140, q=0.0337), and CSF CHIT1 of 5.3% (p=0.0043, q=0.0147). Decreasing CSF levels were found for: CSF pTau217 of −3.0% per year (p=0.0418, q=0.0717), CSF NRGN of −2.5% per year (p=0.0117, q=0.0311) and Aβ40 of −4.0% per year (p<0.0001, q<0.0001).

**Table 2.** Annual percent change and reliability of CSF and plasma biomarkers in AD. Annual biomarker changes were determined using linear mixed effects models. Data was log transformed and models were corrected for age and sex. Intraclass correlation coefficients (ICC) are used as a measure of reliability, values were extracted from the models. The ICC is calculated as a fraction of the between person variability (intercept variance) of the total variance, both between and within person variability (intercept variance + residual variance). ICC values give an indication on the longitudinal stability within one person and lie between 0 and 1. The closer a value is to 1, the lower the within-person variability.

|  | % change per year | p-value change per year | FDR-corrected p-value change per year | ICC |
| --- | --- | --- | --- | --- |
| **CSF – Established markers** |  |  |  |  |
| Aβ42 | -1.5% (-3.9% - 0.9%) | 0.2215 | 0.3127 | 0.83 |
| Aβ40 | **-4.0% (-5.5% - -2.4%)** | **0.0000** | **0.0000** | 0.89 |
| Aβ42/40 | **2.3% (0.2% - 4.6%)** | **0.0373** | **0.0704** | 0.75 |
| tTau | 0.8% (-0.7% - 2.3%) | 0.3117 | 0.4157 | 0.94 |
| pTau181 | -0.2% (-1.7% - 1.3%) | 0.7795 | 0.8134 | 0.95 |
| pTau217 | **-3.0% (-5.8% - -0.1%)** | **0.0418** | **0.0717** | 0.93 |
| NFL | **6.9% (3.7% - 10.2%)** | **0.0000** | **0.0003** | 0.81 |
| YKL40 | **1.4% (0.1% - 2.7%)** | **0.0382** | **0.0704** | 0.95 |
| NRGN | **-2.5% (-4.3% - -0.6%)** | **0.0117** | **0.0311** | 0.93 |
| **CSF – Novel panel markers** |  |  |  |  |
| ABL1 | -2.1% (-6.4% - 2.3%) | 0.3472 | 0.4356 | 0.56 |
| CHIT1 | **5.3% (1.7% - 9.1%)** | **0.0043** | **0.0147** | 0.98 |
| CLEC5A | 0.9% (-1.1% - 3.0%) | 0.3630 | 0.4356 | 0.96 |
| ITGB2 | 1.8% (-0.0% - 3.6%) | **0.0549** | **0.0878** | 0.89 |
| MMP10 | **7.1% (4.4% - 9.9%)** | **0.0000** | **0.0000** | 0.84 |
| SDC4 | **2.2% (0.5% - 3.9%)** | **0.0113** | **0.0311** | 0.82 |
| SPON2 | -0.4% (-2.5% - 1.7%) | 0.6974 | 0.7608 | 0.81 |
| THBD | **4.1% (0.8% - 7.4%)** | **0.0140** | **0.0337** | 0.68 |
| **Plasma markers** |  |  |  |  |
| Aβ42 | 0.3% (-4.4% - 5.2%) | 0.9111 | 0.9111 | 0.40 |
| Aβ40 | 2.4% (-4.1% - 9.3%) | 0.4845 | 0.5538 | 0.11 |
| Aβ42/40 | 1.2% (-0.6% - 2.9%) | 0.1998 | 0.2997 | 0.77 |
| pTau181 | **5.3% (0.6% - 10.4%)** | **0.0291** | **0.0635** | 0.56 |
| pTau217 | **9.8% (3.6% - 16.3%)** | **0.0017** | **0.0082** | 0.66 |
| NFL | **6.6% (2.7% - 10.7%)** | **0.0010** | **0.0058** | 0.70 |
| GFAP | **5.3% (1.7% - 9.0%)** | **0.0042** | **0.0147** | 0.80 |

Among the plasma biomarkers, four of the seven biomarkers changed significantly. Yearly increases over time were found for: plasma pTau181 of 5.3% (p=0.0291, q=0.0635), plasma pTau217 of 9.8% (p=0.0017, q=0.0082), plasma NFL of 6.6% (p=0.0010, q=0.0058), and plasma GFAP of 5.3% (p=0.0042, q=0.0147). The other CSF (Aβ42, tTau, pTau181, ABL1, ITGB2, CLEC5A, and SPON2) and plasma (Aβ40, Aβ42, and Aβ42/40) biomarkers remained stable over time in this cohort. The rate of change in biomarker levels was not significantly affected by clinical stage for these biomarkers. Only on a trend level (p=0.0842) did MMP10 levels increase 4.5% faster in mild AD dementia compared to MCI.

ICC values give an indication on the longitudinal stability within one person and lie between 0 and 1. The closer a value is to 1, the lower the within-person variability. We found that in general ICC values were significantly higher in CSF (mean ICC 0.85) than in plasma (mean ICC 0.57; Table 2). In addition, the more established markers (mean ICC 0.89) had minimally better ICC values compared to the newer biomarkers (mean ICC 0.82), although not significant.

Explorative correlation analysis identified four clusters of biomarkers with correlated rates of change. The biomarkers that correlated together may reflect shared biological processes. Changes in CSF levels of YKL40 correlated with CLEC5A and CHIT1 levels (Supplemental Figure 2). Rates of change in the classical AD biomarkers CSF Aβ40, pTau181, pTau217, tTau, and the synaptic marker NRGN formed a cluster. The third cluster included the following novel CSF markers: THBD, MMP10, SPON2, SDC4, and ITGB2. Plasma pTau181, pTau217, NFL and GFAP also formed a cluster. Results remained similar excluding the participants with CHIT1 SNP variants.

### Sample size calculations

Most clinical trials use a statistical model called an MMRM to assess the treatment response. Therefore, we fitted an MMRM to our placebo biomarker data and used it to estimate the required sample sizes per biomarker to reach either a change between 5-30% or a normalization to the levels of controls of 10-100%. We tested several scenarios because it is expected that different mechanisms of action could have different effects on the biomarker levels. The percent difference between the groups at the end of the trial allows for a more standardized comparison between the different markers. Overall, the plasma biomarkers required approximately 25%-30% bigger sample sizes per group compared to CSF. CSF CHIT1 required very large sample sizes. For the established CSF biomarkers, Aβ42/40 and YKL40 required the lowest sample sizes per arm, n=26 and n=30 respectively for a 20% reduction (Table 3; Figure 2). For the novel biomarker panel, SDC4 (n=14) and ITGB2 (n=26) required the lowest sample sizes per arm. In plasma, Aβ42/40 required the lowest sample sizes, n=21 per arm for a 20% reduction. As a possible biologically relevant scenario we estimated required sample sizes based on % towards normalized or control levels. Biomarkers that reach 100% normalized levels have returned to the levels of controls and are expected to be healthy levels. To reach 30% of the normalized levels the CSF AD biomarkers (Aβ42/40 n=31, tTau n=24, and pTau217 n=11 per arm) markers required the lowest sample sizes. Their plasma counterparts required on average 3 times as many patients per arm. For the novel biomarker panel, ITGB2 showed lowest sample size per arm (n=73), followed by SDC4 (n=78), and MMP10 (n=117). Lastly, we tested what sample sizes are needed to demonstrate stabilization of the disease progression for markers that changes over time (Table 4). To fully stabilize the biomarker levels, CSF MMP10 required the lowest sample sizes per arm (n=181), followed by plasma pTau217 (n=263), and plasma NFL (n=273). To reduce the rate of biomarker change by 50%, all markers needed more than 722 patients per arm.

**Figure 2.**
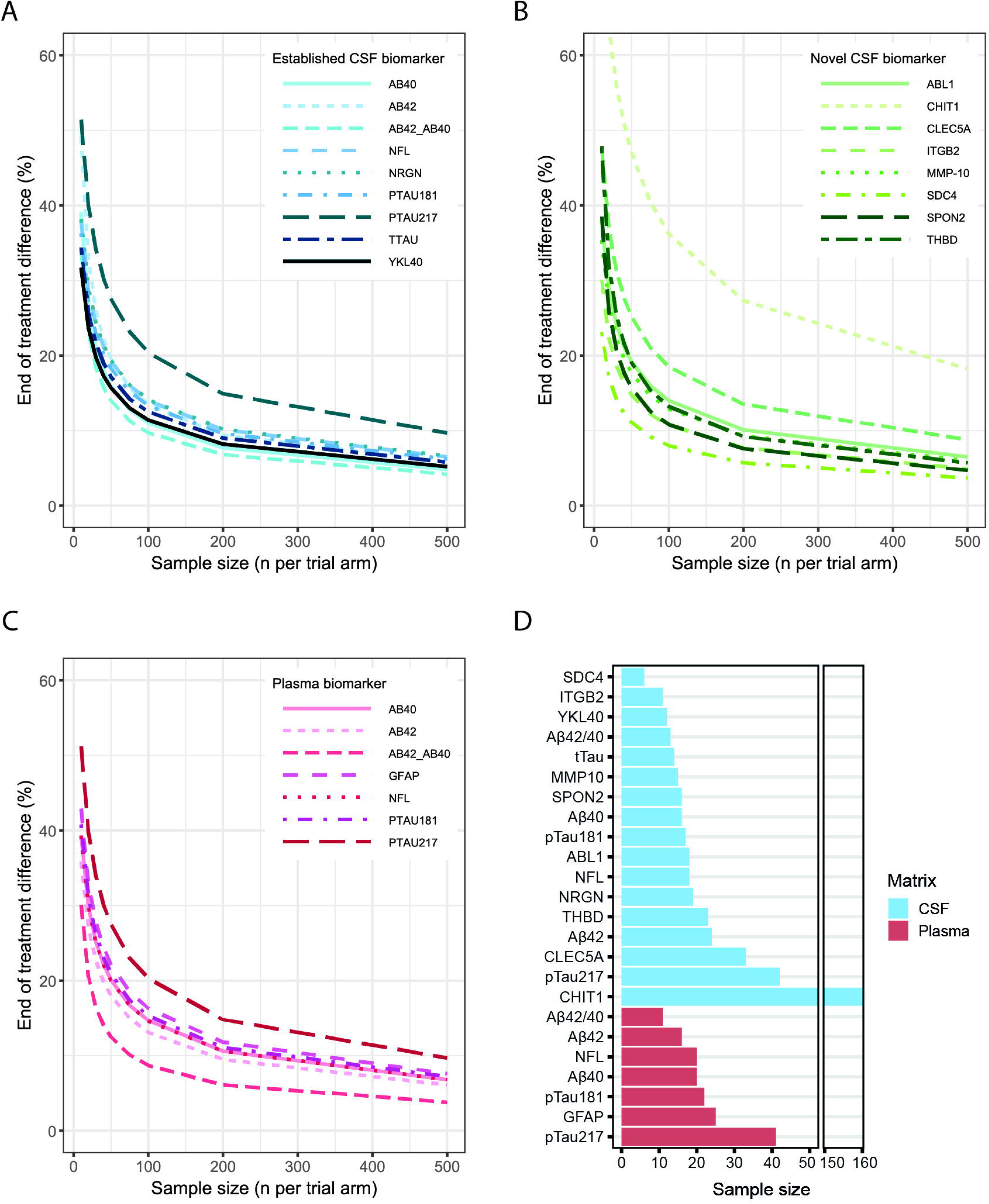
Percent difference at end of treatment per sample size. Estimated percent end of treatment difference for (A) established CSF biomarkers, (B) novel CSF biomarkers, and (C) plasma biomarkers. (D) Overview of the required sample sizes for a 30% treatment effect for all markers

**Table 3.** Estimated required sample sizes per biomarker per effect size. Sample sizes were estimated using the input from the MMRM models as baseline and placebo values. Control levels were derived from the baseline measurements of the current dataset using effect sizes reported on Alzforum and literature. Effect sizes of the expected treatment response were indicated as a percent change from baseline and in relation to control levels. Sample sizes were estimated based on a treatment: placebo ratio of 1:1, 80% power, 20% drop-out rate and significance level of p<0.05. The calculated sample size is indicated as the n per group (treatment or placebo).

|  | 5%<br>difference | 10%<br>difference | 20%<br>difference | 30%<br>difference | %<br>difference<br>control | 10% to<br>normalized<br>levels | 30% to<br>normalized<br>levels | 50% to<br>normalized<br>levels | 100% to<br>normalized<br>levels |
| --- | --- | --- | --- | --- | --- | --- | --- | --- | --- |
| **CSF – Established markers** |  |  |  |  |  |  |  |  |  |
| Aβ42 | 674 | 177 | 49 | 24 | +81.2% | 454 | 51 | 19 | <10 |
| Aβ40 | 459 | 121 | 33 | 16 | +9.5% | 13163 | 1463 | 527 | 132 |
| Aβ42/40 | 361 | 95 | 26 | 13 | +13.5% | 272 | 31 | 11 | <10 |
| tTau | 674 | 160 | 36 | 14 | -59.6% | 216 | 24 | <10 | <10 |
| pTau181 | 775 | 184 | 41 | 17 | -46.4% | 524 | 59 | 21 | <10 |
| pTau217 | 1983 | 470 | 105 | 42 | -90.0% | 91 | 11 | <10 | <10 |
| NFL | 862 | 205 | 46 | 18 | -49.6% | 484 | 54 | 20 | <10 |
| YKL40 | 552 | 131 | 30 | 12 | -26.6% | 1521 | 196 | 61 | 16 |
| NRGN | 882 | 210 | 47 | 19 | -37.8% | 1026 | 114 | 42 | 11 |
| **CSF – Novel panel markers** |  |  |  |  |  |  |  |  |  |
| ABL1 | 867 | 206 | 46 | 18 | -26.5% | 2395 | 267 | 96 | 24 |
| CHIT1 | 7703 | 1826 | 407 | 160 | -49.3% | 4403 | 490 | 177 | 45 |
| CLEC5A | 1596 | 379 | 85 | 33 | -37.7% | 1875 | 209 | 75 | 19 |
| ITGB2 | 487 | 116 | 26 | 11 | -35.8% | 653 | 73 | 27 | <10 |
| MMP10 | 725 | 172 | 39 | 15 | -34.7% | 1047 | 117 | 42 | 11 |
| SDC4 | 263 | 63 | 14 | <10 | -27.0% | 697 | 78 | 28 | <10 |
| SPON2 | 446 | 117 | 32 | 16 | +3.9% | 74139 | 8238 | 2966 | 742 |
| THBD | 645 | 169 | 47 | 23 | +11.7% | 12453 | 1384 | 499 | 125 |
| **Plasma markers** |  |  |  |  |  |  |  |  |  |
| Aβ42 | 752 | 179 | 40 | 16 | -4.7% | 86347 | 9595 | 3454 | 864 |
| Aβ40 | 956 | 227 | 51 | 20 | -4.2% | 135564 | 15063 | 5423 | 1356 |
| Aβ42/40 | 292 | 77 | 21 | 11 | 13.5% | 4326 | 481 | 174 | 44 |
| pTau181 | 1046 | 248 | 56 | 22 | -44.4% | 798 | 89 | 32 | <10 |
| pTau217 | 1958 | 464 | 104 | 41 | -74.2% | 281 | 32 | 12 | <10 |
| NFL | 948 | 225 | 51 | 20 | -46.1% | 655 | 73 | 27 | <10 |
| GFAP | 1197 | 284 | 64 | 25 | -48.1% | 731 | 82 | 30 | <10 |

**Table 4.** Estimated required sample sizes on significantly changing markers. Sample sizes were estimated using the input from the MMRM models as baseline and placebo values. 1.5 year % change was derived from the linear mixed models. Sample sizes were based on no change from baseline or a 50% reduction in the disease progression.

|  | 1.5 year % change | No change from baseline | 50% reduction |
| --- | --- | --- | --- |
| **CSF markers** |  |  |  |
| Aβ40 | -5.9% | 297 | 1187 |
| NFL | 10.5% | 227 | 905 |
| CHIT1 | 8.1% | 3353 | 13410 |
| MMP10 | 10.8% | 181 | 722 |
| SDC4 | 3.3% | 652 | 2607 |
| **Plasma markers** |  |  |  |
| pTau181 plasma | 8.1% | 451 | 1803 |
| pTau217 plasma | 15.0% | 263 | 1052 |
| NFL plasma | 10.0% | 273 | 1089 |
| GFAP plasma | 8.0% | 533 | 2131 |

### Estimating the effect of heterogeneity on effect sizes in a clinical trial

The data was used to simulate the biomarker results of hypothetical clinical trials. Even without any treatment effect, random chance can cause the groups to differ if patients with high rates of change or low rates of change in biomarker levels are not equally distributed among the placebo and treatment group. When there are more patients with a high rate of change in the placebo group, the biomarkers could change significantly more from baseline in the placebo group compared to the treatment group, which makes it appear as though there is a positive treatment effect. However, this is not representative of a true treatment effect. The simulations provided insights into how this random oversampling could affect the treatment effect and trial outcome and the 95% CI provided an estimate of the possible range of false positives. Figure 3A shows the 95% CIs of the group differences for a hypothetical phase 2B trial (n=200) in the absence of a treatment effect. Overall, the 95% CIs were wider for plasma (average 9.4% ranging from 4% to 15%) compared to CSF (average 5%, ranging from 3% to 9%) biomarkers, meaning that plasma markers may be more sensitive to non-perfect randomization, and could yield false-positive results at higher effects sizes compared to CSF. In CSF, the marker showing the lowest 95% CI, meaning the effect of heterogeneity is smallest, was YKL40 (3%). The largest interval was shown for CSF ABL1 (9%). In plasma, the marker with the smallest interval was the Aβ42/40 ratio (4%) and the largest interval was that of plasma pTau217 (15%). Generally, the biomarkers that had high ICC values also showed smaller 95% CIs. The outcome of the simulations were similar for both clinical groups (Figure 1B). Lastly, we showed how increasing the sample sizes reduced the 95% CI, for example the 95% CI for plasma pTau217 for n=50 is ~30%, for n=200 ~15%, and for n=2000 ~5%, to quantify how increasing sample sizes reduced the effects of heterogeneity on the outcomes (Figure 3B).

**Figure 3.**
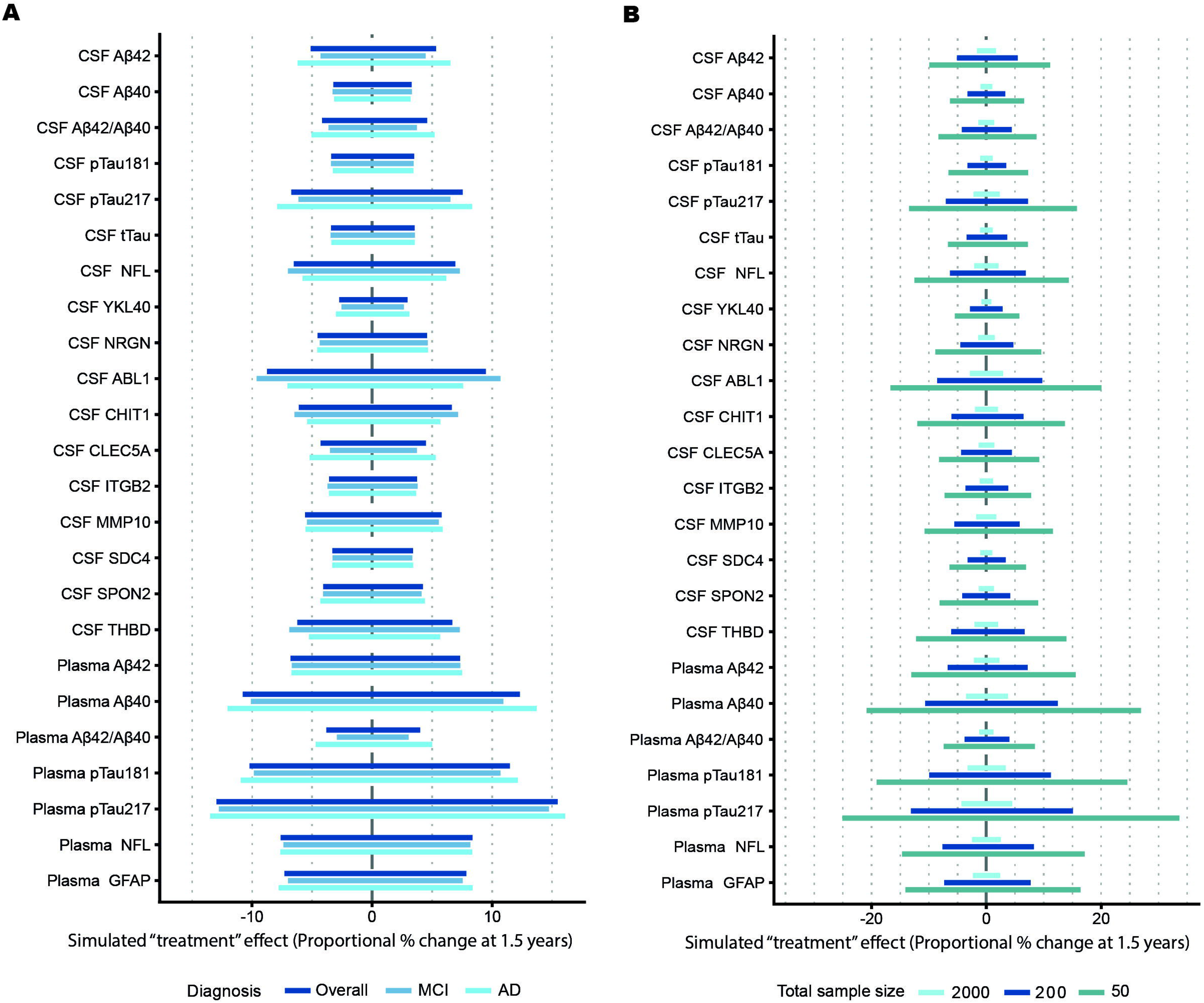
Effects of heterogeneity in disease progression on clinical trial outcomes. (A) The simulated 95% confidence intervals of the “treatment” effects of a clinical trial (n=200) including a mix of MCI due to AD and mild AD dementia, only MCI due to AD or only AD dementia. (B) The effects of different sample sizes on the 95% confidence intervals of the trial simulation.

## Discussion

Despite the importance of biological readouts for early AD trials, the natural trajectories for a broad range of markers within the duration of a trial are not well established. We used data from a large phase 2 study to measure CSF and plasma biomarker trajectories within a clinical trial setting, and investigate how within- and between-person variation in rates of change can affect the power and treatment response reliability. The majority of the biomarkers showed annual changes that were consistent with increasing disease severity. Counterintuitively, a selection of markers showed changes towards normalization. To demonstrate a 30% treatment effect at end of treatment, similar to the biomarker effects reported with lecanemab and donanemab treatment, almost all markers required less than 45 patients per treatment group. Plasma biomarkers showed more variability in rates of change between individuals and are therefore predicted to be more sensitive to non-perfect randomization. Increased sample sizes mitigated the effects of heterogeneity in rates of change. Clinical severity, MCI versus mild dementia, showed minimal effect on the observed heterogeneity, supports the validity of combining these clinical groups in standard clinical trial practice. The findings form a frame of reference for the interpretation of clinical trial results and provide practical estimates to guide study design.

CSF NFL, YKL40, SDC4, MMP10, CHIT1, and plasma NFL, GFAP, pTau181, and pTau217 all increased over time, which is suggestive that these markers track disease progression, in line with cross-sectional of AD^1,26,30,56–63^. Only MMP10 levels showed, at a trend level, faster increases in mild dementia stage compared to MCI stage. Counterintuitively, for a selection of the markers (CSF pTau217, NRGN, the Aβ42/40 ratio, and CSF THBD), the annual rates of change were suggestive of an improvement. Previous research on the established AD biomarkers has shown that in the dementia stage, Aβ40, pTau and NRGN decrease longitudinally, suggesting this is a common end stage phenomenon^24,26,31,64^. For THBD, this seems the case too, and in general the ICC and small changes compared to controls suggest that it is not promising outcome marker in the context of AD^1^. For the biomarkers that change towards normal levels, it is crucial a placebo group is added to the clinical trial to warrant correct interpretation when they are employed as an endpoint.

Interestingly, CSF rates of changes in pTau217 and NRGN clustered together with CSF Aβ40, tTau, and pTau181, forming an overall AD related biomarker cluster. Rates of change from CSF CHIT1, YKL40 and CLEC5A also clustered together. All three of these markers are expressed by macrophages and microglia and have been linked to innate immunity and white matter involvement in AD^42,43,65–69^. Lastly, the changes in CSF THBD, MMP10, SPON2, SDC4, and ITGB2 clustered together. This third cluster seems to be a mix of proteins related to blood-brain barrier function and angiogenesis^41,70–79^. There is a big gap in the use of biomarkers in drugs targeting inflammation and vasculature^3,80^. Our exploratory correlation analyses suggest that proteins involved in inflammation and vasculature change during natural disease progression. A combination of biomarkers within such a cluster could be a good biomarker panel to explore if there are consistent treatment effects on the respective biological processes. It is also foreseeable that the biomarker with the greatest effect size and good technical characteristics can be selected as primary endpoint as a proxy of the target. In a broader context, it shows how studying longitudinal biomarkers trajectories simultaneous with drug development may help to reduce the gaps in biomarkers for trials targeting these mechanisms^3^.

We showed that the required sample sizes for plasma were approximately 25-30% higher compared to their CSF counterparts. This is not surprising as there is more interference of the periphery in plasma and comorbidities can have a bigger effect on plasma biomarkers. However, it is also easier to recruit patients and conduct trials without lumbar punctures, meaning that the optimum will depend on the unique information that can be collected from CSF. The simulations provided estimates of end of treatment group differences when there was no treatment effect, which means that these differences were likely caused by variation in biomarker trajectory. CSF biomarkers (average 5% “treatment” effect) outperformed plasma biomarkers (10% “treatment” effect), which indicated that it is less likely to find false positives in CSF compared to plasma at the same effect sizes. We also clearly showed how increasing the sample size reduced this heterogeneity effect.

There are several strengths and limitations to this study. The use of CSF and plasma biomarkers in clinical trials for AD has been widely acknowledged^85–87^. Firstly, while our set-up provided results that are applicable in a clinical trial setting, it could not capture the whole disease trajectory. To study that, follow-up times are ideally longer and a population representative of all AD patients is included. In addition, to distinguish the effect of AD from possible effect of general aging, a healthy control group is needed. Secondly, although the required sample size estimates were designed to be as translatable as possible, assumptions regarding power, drop-out, and analyses may limit generalizability. By covering a range of estimated effect sizes, we aimed to provide useful required sample sizes applicable to multiple future trials. The biomarkers were measured by a range of different assay methods, although all were immuno-based, we could not exclude the possibility that other assay brands or next-gen versions would yield a different power. Still, the measurements adhered to clinical chemistry standards, therefore, large deviations are not expected. Strengths included that the current trial is representative in terms of inclusion criteria, duration, the biomarkers measured, and that we employ commonly used statistical models as the basis for sample size calculations to increase translatability to future trials. The included trajectory analyses and insights on overall variability and stability could also be used as reference and starting point for future clinical trial design in mild AD. It would be of value to repeat such analyses for the biomarker effects in a representative placebo group of the even earlier asymptomatic stages of AD^85,88^.

The current study aimed to provide guidance for future clinical trial design. The biomarker trajectories provided insights into general biomarker changes over 1.5 years and could be used as a comparison to placebo data from smaller clinical trials. Moreover, we showed that generally biomarkers are stable within persons, increasing their applicability in a clinical trial setting and simplifying the interpretation. The required sample size estimates could be used to determine sample sizes in future clinical trials. Lastly, the trial simulations offered insights into the effects of heterogeneity in biomarkers changes and how these might affect biomarker outcomes and trial interpretations.

## Data sharing

This clinical trial dataset is not publicly available. Qualified researchers can apply for collaboration projects and federated analyses.

## Author contributions

MO, LV, AL, CET designed the study. MO, MWTT, AL, LV, BMT, RJJ aided in/performed the statistical analyses. YSH, LiV, AA prepared samples for biomarker analyses. TO, PvB PS, EGBV designed and contributed to the clinical trial. MO wrote first draft of the manuscript. AL drafted the figures. LV and CET were major contributors in writing the manuscript. All authors read and contributed to the revisions and editing of this manuscript.

## Acknowledgements

NA

## Figure titles and legends

Sup. Figure 1 Biomarker trajectories over 1.5 years.

Legend: Biomarker trajectories are shown analyzed with the linear mixed models (blue lines) including individual patient values (grey lines) and analyzed with the mixed model for repeated measures (black lines).

Sup. Figure 2 Correlation heatmap on the rate of change.

Legend: Pearson correlation between the rates of change of all biomarkers. P values were FDR corrected. * p<0.05, ** p<0.01, *** P<0.001

