## Supplemental Figure 1 for "Treatment response biomarkers in early Alzheimer’s disease: longitudinal trajectories, sample size estimates, and the impact of progression variability"

### Slide 1
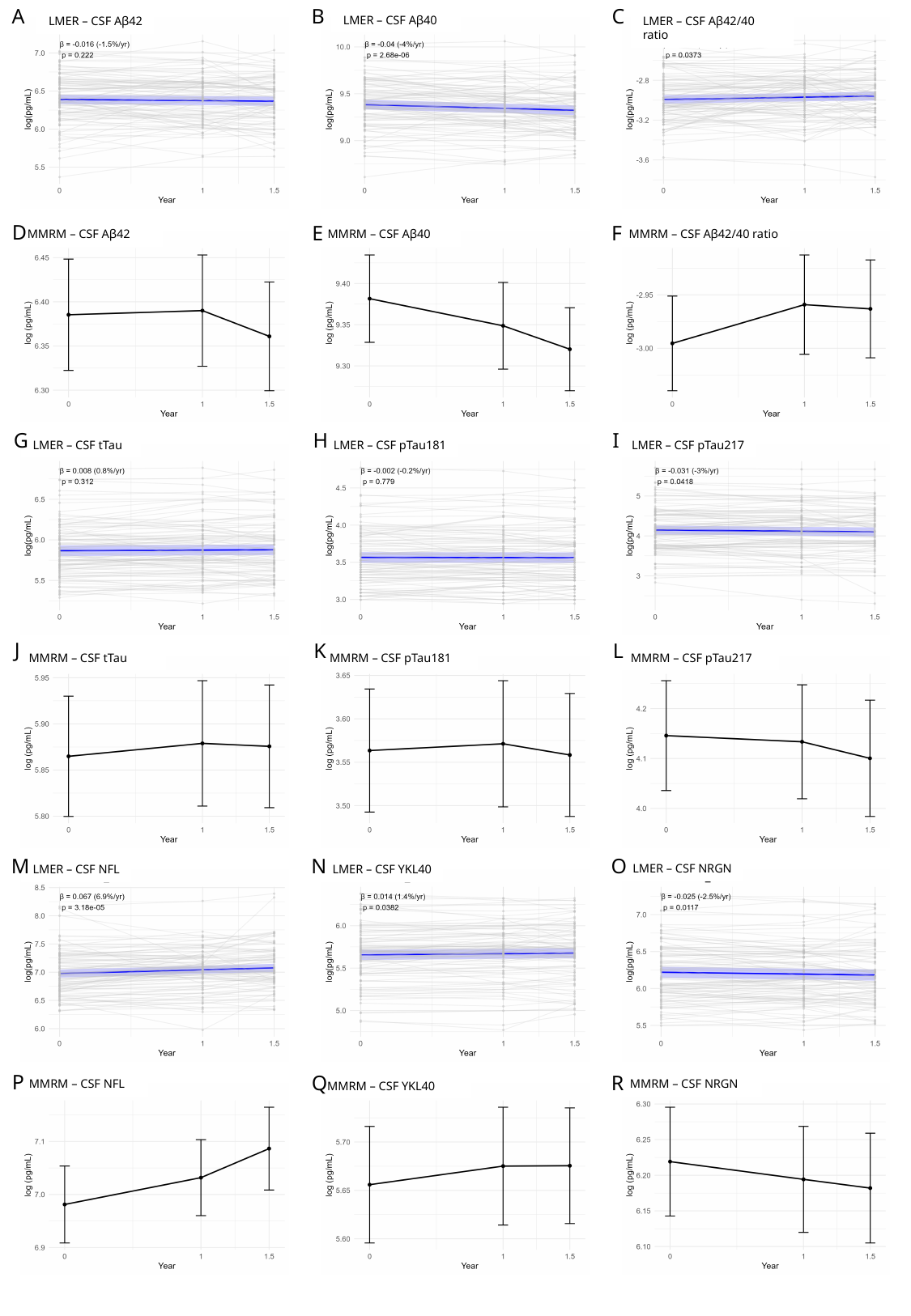

B
C
A
LMER – CSF Aβ40
LMER – CSF Aβ42/40 ratio
LMER – CSF Aβ42
D
E
F
MMRM – CSF Aβ42
MMRM – CSF Aβ40
MMRM – CSF Aβ42/40 ratio
H
I
G
LMER – CSF tTau
LMER – CSF pTau181
LMER – CSF pTau217
J
K
L
MMRM – CSF tTau
MMRM – CSF pTau181
MMRM – CSF pTau217
N
O
M
LMER – CSF NRGN
LMER – CSF NFL
LMER – CSF YKL40
P
Q
R
MMRM – CSF NFL
MMRM – CSF NRGN
MMRM – CSF YKL40

### Slide 2
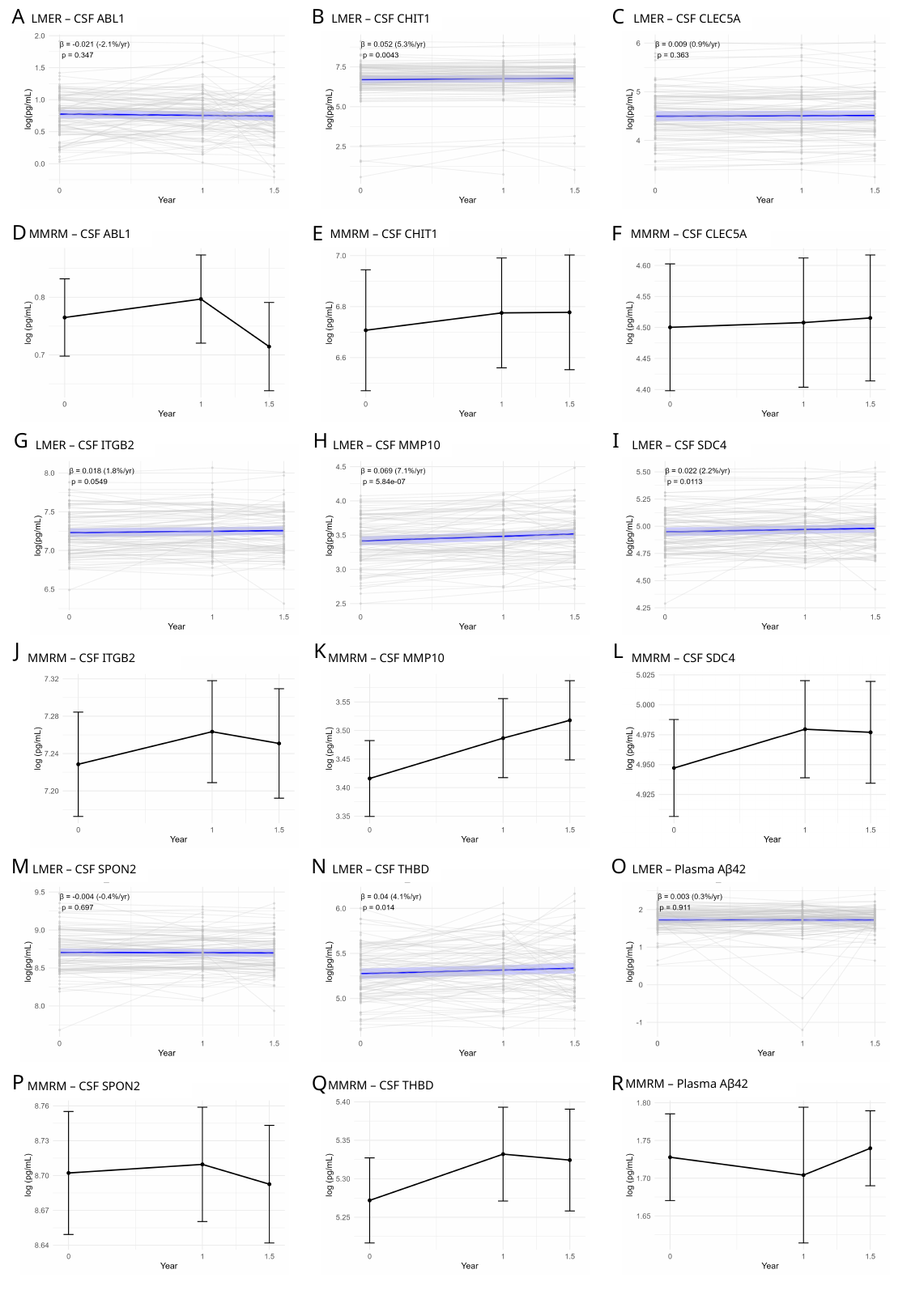

B
C
A
LMER – CSF ABL1
LMER – CSF CHIT1
LMER – CSF CLEC5A
D
E
F
MMRM – CSF ABL1
MMRM – CSF CHIT1
MMRM – CSF CLEC5A
H
I
G
LMER – CSF ITGB2
LMER – CSF MMP10
LMER – CSF SDC4
J
K
L
MMRM – CSF ITGB2
MMRM – CSF MMP10
MMRM – CSF SDC4
N
O
M
LMER – CSF SPON2
LMER – CSF THBD
LMER – Plasma Aβ42
P
Q
R
MMRM – Plasma Aβ42
MMRM – CSF THBD
MMRM – CSF SPON2

### Slide 3
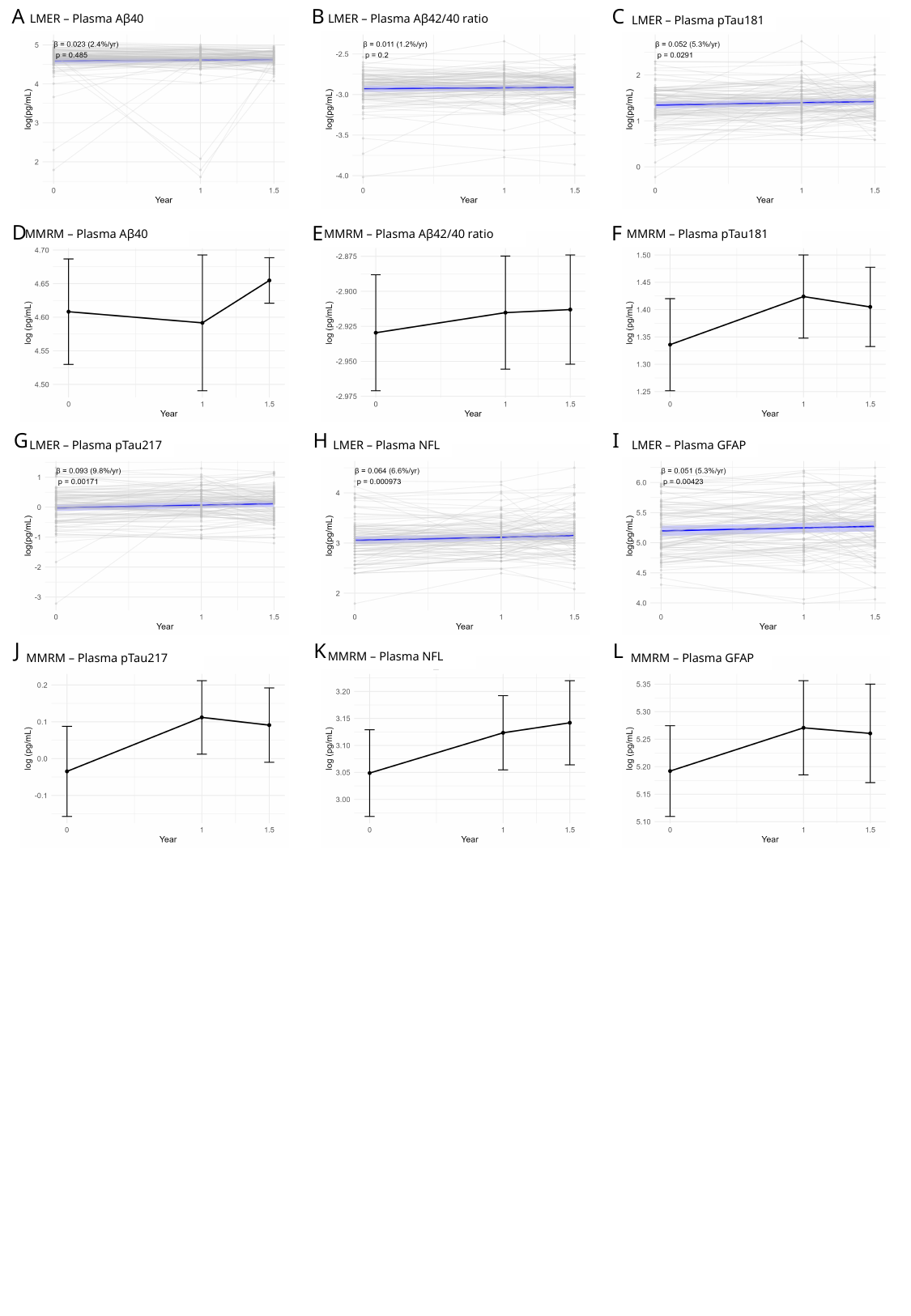

B
C
A
LMER – Plasma Aβ40
LMER – Plasma Aβ42/40 ratio
LMER – Plasma pTau181
D
E
F
MMRM – Plasma Aβ40
MMRM – Plasma Aβ42/40 ratio
MMRM – Plasma pTau181
H
I
G
LMER – Plasma pTau217
LMER – Plasma NFL
LMER – Plasma GFAP
J
K
L
MMRM – Plasma NFL
MMRM – Plasma pTau217
MMRM – Plasma GFAP
