## Supplementary figures and images for "Treatment response biomarkers in early Alzheimer’s disease: longitudinal trajectories, sample size estimates, and the impact of progression variability"

### Supplemental Figure 2

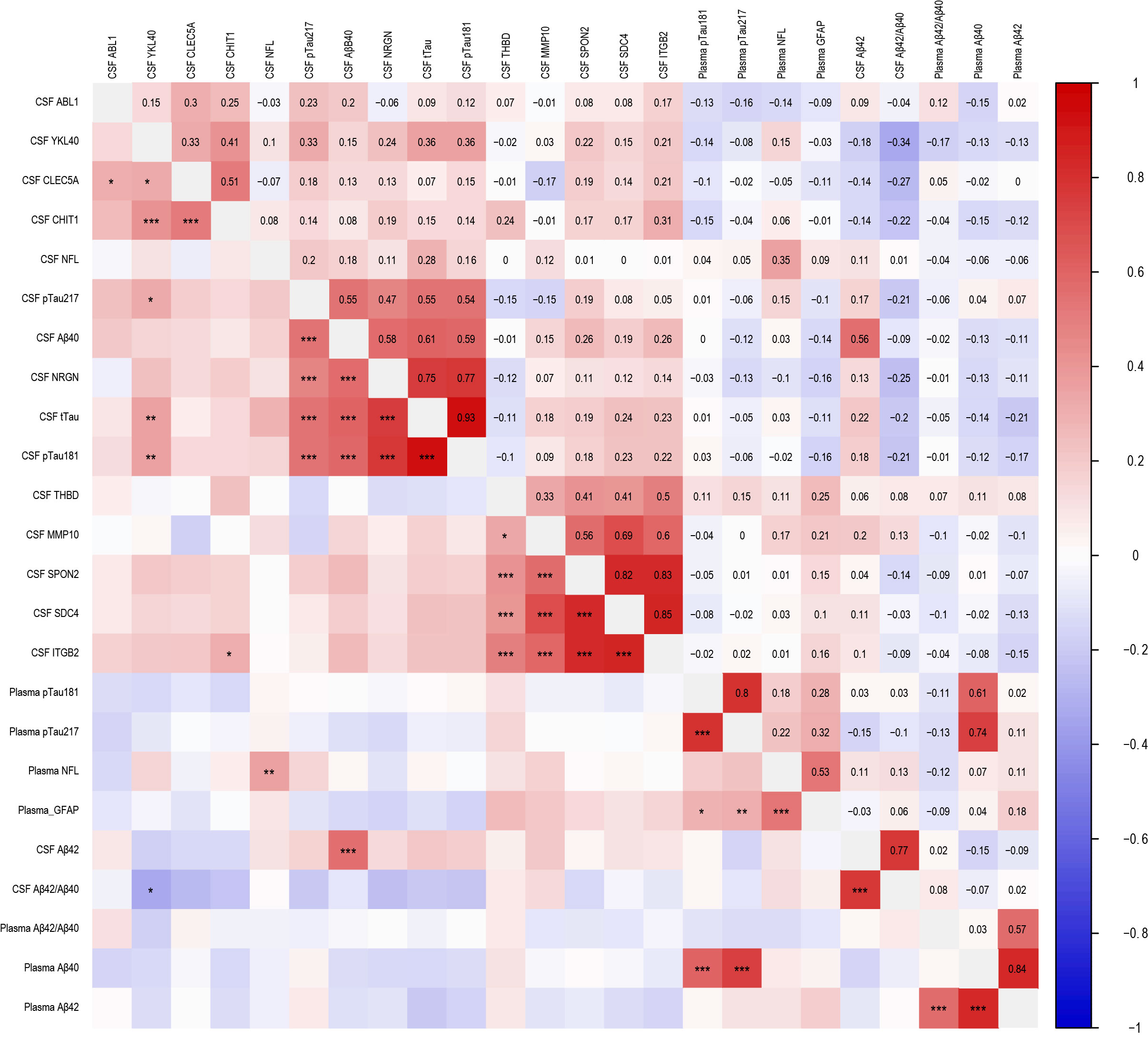
